# MostCare-Based Assessment of Cardiac cycle efficiency in Coronary Artery Disease Patients: High-flow nasal cannula versus standard oxygenation for gastrointestinal endoscopy with sedation. The prospective single-center randomised controlled MEHIS study protocol

**DOI:** 10.1101/2024.07.27.24311111

**Authors:** Fang Xie, Mu Jin, Tingting Ma, Xiaorui Zhou, Sheng Wang

## Abstract

**Introduction:** During gastrointestinal endoscopy (GIE) procedures(upper/lower) performed under deep sedation, patients with coronary artery disease(CAD) have poorer tolerance, with higher incidence of hypotension and myocardial ischemia. Patients with CAD should particularly avoid hypoxemia caused by deep sedation and increased oxygen consumption caused by inadequate sedation. Recent data indicate that high-flow nasal oxygen therapy (HFNO) is recommended for preventing hypoxemia in high-risk patients. The data on cardiac cycle efficiency (CCE) from MostCare can be used to assess myocardial oxygen supply-demand balance in patients with CAD. HFNO may potentially improve myocardial oxygen supply during GIE under deep sedation. We hypothesize that compared to standard oxygen therapy (SOT), HFNO could improve CCE in patients with CAD.

**Methods and analysis:** The MEHIS (MostCare-Based Assessment of CCE in CAD Patients: HFNO versus SOT for GIE with sedation. The prospective single-center randomised controlled) study is a single-center randomized controlled trial comparing the effects of HFNO and SOT during GIE under deep sedation administered by anaesthesiologists in the procedure room in patients with CAD. Ninety patients will be randomly allocated in a 1:1 ratio to two parallel groups. The primary outcome is the difference in CCE levels between the two groups during sedation. Secondary outcomes are the incidence of hypotension(hypotension defined as a systolic blood pressure below 80 mmHg), values of BNP (brain natriuretic peptide), TnI (troponin I), and lactate levels at 6-12 hours post-operation,the occurrence of hypoxemia defined as SpO2 measurement equal to or below 92%, MostCare hemodynamic parameters excluding the primary outcome, interventions required to maintain upper airway patency, patient agitation episodes (assessed by touching the oxygen supply device), and presence of intraoperative adverse memories postoperatively.

**Trial registration number:** ChiCTR2400086887

**Strengths and limitations of this study:** This is the first pragmatic randomized single-center study comparing HFNO to SOT for oxygenating patients with CAD undergoing GIE.

In contrast to comparing HFNO with high fraction of inspired oxygen (FiO_2_) typically used in SOT with low oxygen flow and consequently lower FiO2, this study adjusts gas flows to target roughly the same level of applied FiO2 in both groups. This approach aims to assess whether HFNO can improve CCE through positive end-expiratory pressure and/or dead space washout effects.

In SOT group, precise FiO2 cannot be guaranteed. That is why we utilized a pre-existing abacus to achieve the best equivalence.

Technically, blinding of practitioners and other nursing staff to the study groups is not feasible. However, the printout of recorded primary outcome measures is produced, allowing delayed reading by assessors blinded to treatment allocation. We employed a blind method in the study of patients.

In patients not undergoing endotracheal mechanical ventilation, MostCare data collection may exhibit slight bias due to respiratory influences. However, both groups in our study were conducted under sedation, and longitudinal comparisons were performed, thus minimizing the impact of such bias.

CAD patients with upper/lower gastrointestinal bleeding often suffer from anemia. We stratified CAD patients into anemic and non-anemic groups and compared the CCE under two oxygen delivery modes. This could provide a basis for further evaluating the oxygen therapy effectiveness of HFNO for anemic patients.

## Introduction

Sedation during GIE improves the quality of examination, patient comfort and allows performing complex procedures.^1^ CAD patients mostly take anticoagulant medications such as aspirin and clopidogrel, etc., which often lead to gastrointestinal bleeding,consequently increasing the proportion of GIE. Due to the presence of underlying heart disease and multiple comorbidities, CAD patients have poor tolerance to anesthesia. Hypoxia and circulatory fluctuations occurring during sedation often lead to myocardial ischemia. The recent American Society for Gastrointestinal Endoscopy (ASGE) guidelines suggest that sedation, under the responsibility of anaesthesiologists, should be considered for patients with multiple medical comorbidities including CAD patients.^2^

During the process of GIE under deep sedation, myocardial ischemia caused by hypoxemia and hypotension is the most common complication in patients with CAD. CAD patients often have comorbid obesity. Sedation-induced myorelaxation and respiratory depression may cause upper airway obstruction and a decrease in functional residual capacity, thereby exacerbating hypoxemia. Patients taking anticoagulant medications may experience upper or lower gastrointestinal bleeding, leading to anemia, which causes a rightward shift of the oxygen dissociation curve and decreased oxygen affinity. Additionally, CAD patients with concomitant hypertension often have poor vascular elasticity, making them prone to hypotension during sedation, further reducing organ tissue perfusion. Hypoxemia and hypotension may lead to myocardial ischemia, arrhythmias, and cerebral hypoxia, causing adverse cardiovascular and cerebrovascular events, thereby increasing mortality and hospitalization time. Therefore, CAD patients should ensure the balance of myocardial oxygen supply and demand, stable hemodynamics to avoid myocardial ischemia.

In recent years, an increasing number of studies have begun to focus on optimizing the hemodynamics of CAD patients. Although pulmonary artery catheterization (PAC) has been the gold standard for assessing overall cardiac status for nearly fifty years, it is associated with a high incidence of complications and complex procedures. The FloTrac system is less invasive and provides continuous cardiac output (CO) monitoring. However, its accuracy is limited in unstable patients, severe arrhythmias, severe aortic valve regurgitation, and in patients with other factors that interfere with arterial waveform.^3^ Comparable to PAC and PiCCO,MostCare demonstrates excellent consistency not only in hemodynamically stable patients^4^,but also in critically ill patients with hemodynamic instability. In pediatric patients undergoing cardiac surgery, MostCare demonstrates consistency with Fick method measurements ^5^ and transesophageal Doppler echocardiography^6^ in assessing overall cardiac function.^7^ After cardiac surgery, infants entering the ICU, MostCare data can effectively monitor cardiac dysfunction and predict mechanical ventilation duration.^8^ In adult patients undergoing off-pump coronary artery bypass surgery and those receiving veno-venous extracorporeal membrane oxygenation therapy, the use of MostCare shows good consistency and trending in detecting cardiac output compared to intermittent pulmonary artery thermodilution^9^ and transthoracic echocardiography^10^, respectively. Furthermore, in hemodynamically unstable patients (including those with conditions such as ventricular failure, vasoplegic shock, hypertensive crisis, hypovolemic shock, and aortic valve stenosis), there exists a good correlation between MostCare data and high-fidelity human patient simulators. ^11^ In septic patients, when the arterial waveform is accurate, MostCare and PiCCO transpulmonary thermodilution exhibit good agreement even after the reduction of norepinephrine and changes in vascular tone or volume expansion. ^12^ Despite controversies, when MostCare is used for unstable patients reliant on high doses of positive inotropic agents or even IABP patients with sinus rhythm, its monitoring data aligns with pulmonary artery catheterization data,^13^ unaffected by the inflation and deflation of the intra-aortic balloon pump. ^14^ MostCare data can also demonstrate good predictability of hypotension during the induction of anesthesia, identifying patients at risk of anesthesia-induced hypotension.^15^ Therefore, MostCare can be used to monitor and optimize the hemodynamics of CAD patients undergoing non-intubated sedation for gastrointestinal endoscopy.

MostCare monitoring data of CCE is a comprehensive indicator reflecting the venous system, cardiopulmonary interaction, and coupling status between the ventricle and arterial system (V-A Coupling), which can effectively indicate whether the left ventricular function is optimized. Real-time monitoring of perioperative left ventricular function using CCE, allowing timely intervention, may play a crucial role in maintaining cardiopulmonary function and promoting rapid postoperative recovery in patients. CCE calculates the ratio of cardiac systolic energy consumption to total heartbeat energy consumption from the perspective of pressure waveform energy. An increase of CCE reflects a decrease in the energy required to maintain the same hemodynamic balance within the cardiovascular system,which can be interpreted as an improvement in ventricular-arterial coupling. ^16^ CCE can effectively assess cardiac circulatory function not only in post-cardiac surgery patients undergoing mechanical ventilation,^17^ but also in hospitalized patients,^18^ pediatric ICU patients^19,20^ and non-intubated patients^21,22^. During the first 48 hours following pediatric cardiopulmonary bypass, CCE is the most sensitive indicator for assessing improvements in systemic hemodynamics and myocardial performance.^23^ A six-month follow-up study found that the occurrence of adverse events was directly proportional to NT-proBNP levels and inversely proportional to CCE, suggesting that CCE can identify high-risk patients following cardiac surgery.^24^ Furthermore, in non-mechanically ventilated patients, although five minutes of negative pressure ventilation can significantly improve CCE in healthy subjects,^16^ its impact on extubated patients after cardiac surgery is minimal.^21^ This highlights the assessment value of CCE in non-intubated sedated patients with CAD. More importantly, CCE can not only assess the overall cardiac function of patients with severe aortic valve stenosis undergoing TAVI surgery under deep sedation and femoral nerve block anesthesia^22^ but also determine whether there is an improvement in cardiac function after TAVI surgery.^25^ Therefore, CCE may have indicative significance for the occurrence of myocardial ischemia in non-intubated sedated patients undergoing GIE with CAD.

To prevent myocardial ischemia in patients with CAD, the American Society of Anesthesiologists and the American Society for Gastrointestinal Endoscopy recommend the use of oxygen therapy.^2^ SOT is a widely used method at present. However, HFNO can provide higher oxygen flow rates and more precise fraction of inspired oxygen (FiO2)^26^, potentially offering better assessment of CCE and myocardial protection. The gas is heated, humidified, and delivered through large-bore nasal cannulas, enhancing patient comfort and tolerance. The gas flow rate is typically set between 30–70 L/min, allowing for the delivery of accurately known fractions of inspired oxygen (FiO2) ranging from 21% to 100%. High gas flow creates resistance against expiration, maintaining mild positive end-expiratory pressure (PEEP) in the airways, while also generating a dead space washout effect.^27^ By maintaining good airway patency and respiratory function, HFNO may reduce myocardial workload and decrease cardiac burden. Additionally, the PEEP effect of HFNO helps improve alveolar ventilation, alleviate pulmonary edema, thereby reducing right ventricular load and promoting coronary artery perfusion.

HFNO has many clinical applications in the intensive care unit (ICU) and the operating room (OR),^28^ but its utility for procedural sedation is still underestimated. In the neuroanesthesia practice and electroconvulsive therapy, HFNO can improve arterial oxygenation by providing higher inspiratory oxygen concentration and maintaining higher dynamic positive airway pressure.^29^ In patients undergoing oral maxillofacial surgery and/or dental treatment, HFNO can maintain oxygenation and possibly prevent hypercapnia.^30^ In elderly patients undergoing endoscopic retrograde cholangiopancreatography (ERCP),HFNO may provide adequate oxygenation without causing procedural interruption under deep sedation.^31^ During pediatric procedural sedation in congenital heart disease, HFNC could reduce the incidence of desaturation, the need for airway assisted ventilation and risk of carbon dioxide retention without causing hemodynamic instability or gastric distention.^32^ HFNO has been shown to reduce the risk of desaturation in adults receiving procedural sedation and analgesia during AF ablation. ^33^ During transfemoral transcatheter aortic valve replacement, there was a lower incidence of oxygen desaturation and a significantly higher comfort score in the HFNO group. Anyway, HFNC may reduce the incidence of hypoxemia in patients at moderate to high risk for hypoxemia.^34^

To date, there have been no studies conducted on the use of HFNO in CAD patients under deep sedation, either for upper gastrointestinal endoscopy or lower. Furthermore, there have been significant variations in the settings of HFNO in previous studies: typically comparing FiO2 set at 100% with standard low-flow oxygen, and the flow rates of HFNO used vary across different studies as well. Most importantly, it remains unclear whether HFNO can improve CCE and consequently reduce the incidence of myocardial ischemia in patients with CAD.

Hence, we hypothesize that HFNO during GIE under sedation could decrease the incidence of myocardial ischemia in comparison with SOT by decreasing hypoxemia, circulatory fluctuations, and improving CCE. For this purpose, changes in cardiac enzymes were monitored to comprehensively evaluate the myocardial protective effects of HFNO.

## Methods and analysis

### Design

This is an investigator-initiated, prospective, single-center, randomized controlled, superiority trial comparing the efficacy HFNO with SOT in CAD patients undergoing GIE performed under sedation. Our primary hypothesis is that compared to SOT, HFNO may improve CCE, reduce the incidence of hypotension and lower postoperative myocardial enzymes. Ninety eligible patients will be randomly assigned in a 1:1 ratio to two parallel groups, with 45 patients in each group.

A computer-generated randomisation is performed with stratification in a 1:1 ratio, using a web-based management system. After randomisation, the intervention (HFNO or SOT) is initiated. The randomly assigned groups will be documented in patient medical records and dedicated charts, which will summarize all the patient populations allocated throughout the trial.

Oral and written information will be provided during the anaesthesia consultation several days before endoscopy. Written consent will be obtained on the day of the GIE procedure after eligibility verification. Patients have the right to withdraw their consent and terminate their participation at any time for any reason. We used the SPIRIT reporting guidelines in our study.^35^

Patients are expected to be included during a 12-month period starting in July 2024.

- July 2024: Ethical approval for the protocol and development of trial tools (case report forms, randomization system).
- August 2024 -May 2025: Patient enrollment.
- June 2025: Database cleaning and closure. Data analysis, manuscript writing, and submission to peer-reviewed journals for publication.

### Study setting

The trial is conducted at Beijing Anzhen Hospital. A senior anesthesiologist conducted anesthesia sedation procedures before and after the research. At the same time, data recording and collection were carried out by an anesthesiologist.

### Study objectives

The main research objective is to compare the CCE. CCE is collected and generated by MostCare, recording values before and after sedation, every 3 minutes after induction, and after patient awakening.

The secondary parameters are to compare between groups:

- The incidence of hypotension. Hypotension is defined as a systolic blood pressure below 80 mmHg.
- Postoperative BNP, TnI, and lactate levels at 6-12 hours between the two groups.
- The incidence of hypoxemia (defined as SpO2 ≤92%).
- The frequency of patient agitation episodes (defined as touching the oxygen supply device).
- Intraoperative adverse memory recall.
- The need for mask ventilation or any airway intervention.
- The duration of sedation (from the induction to the awakening of the patient).

The secondary hemodynamic parameters are to compare between groups before and after sedation, every 3 minutes after induction, and after patient awakening:

- Heart rate (HR).
- Systolic blood pressure, diastolic blood pressure and mean arterial pressure, respectively (SBP, DBP and MAP).
- Pdic-a.
- Stroke volume index (SVI).
- Systemic vascular resistance index (SVRI).
- Cardiac index (CI).
- Pulse pressure variation (PPV).
- The maximum rate of pressure increases in the left ventricle during systole (dP/dtmax).

### Data collection

The monitoring data recorded by MostCare and SpO2 are automatically conducted. As in the routine clinical setup, the standard arterial pressure transducer is connected to the Mindray monitor (Mindray, BeneVision N15 OR) via a dual-output pressure module. For the study, the pressure module was also directly connected to the Mostcare device(Bio-Si International, italy) to allow continuous transmission of the original signal and sampling at 1000 Hz. **Mostcare** provides averaged beat-to-beat calculated data within 30 seconds, including pressure, and continuously displays the data on the screen. For each parameter, detailed 2-minute measurements at 30-second intervals recorded by Mostcare were downloaded to Excel sheets for offline analysis. Subsequently, the four consecutive measurements were averaged. During the intervention, SpO2 is recorded every minute using a Mindray monitor (Mindray, BeneVision N15 OR) to capture hypoxemia defined as SpO2 ≤ 92%. Outcomes are recorded on paper case report forms (CRF). Data collection stops when the patient leaves the recovery room. The trend of vital signs (including SpO2) and Mostcare hemodynamic records are printed out and placed into an envelope with the CRF.

Investigators not involved in patient care and blinded to the allocated intervention will read and review the recorded data to check for consistency with the events reported by clinicians on the paper CRF.

### Sample size

In a short preliminary observational study of 20 CAD patients undergoing GIE with the usual SOT with sedation, we found a 0.2 decrease of CCE compared to baseline values. Based on published studies showing a 0.1 increase in CCE with different oxygenation devices compared with standard oxygenation^16^, we hypothesize that HFNO will reduce the decrease in CCE from 0.2 to 0.1. With α risk set at 5% on both sides and a statistical power of 80%, 37 patients per group are required to demonstrate such a difference. We chose to increase this number to 45 patients per group to account for potential dropouts for any reasons and possible unreliable CCE readings identified by the anesthesia team due to technical issues.

### Recruitment and informed consent

The researchers provided the patients with the information letter and explanations of the study are given to the patient one day before the surgery (see online supplementary file 1).

The randomisation is performed at the entrance in the operating room and arterial puncture procedure were conducted by the researcher. The patients who accept to participate sign the written informed consent form at the same time (see online supplementary file 2).

Figure1 shows the research protocol execution process. Figure 2 shows the schedule of enrolment, interventions and assessments.

**Fig 1:**
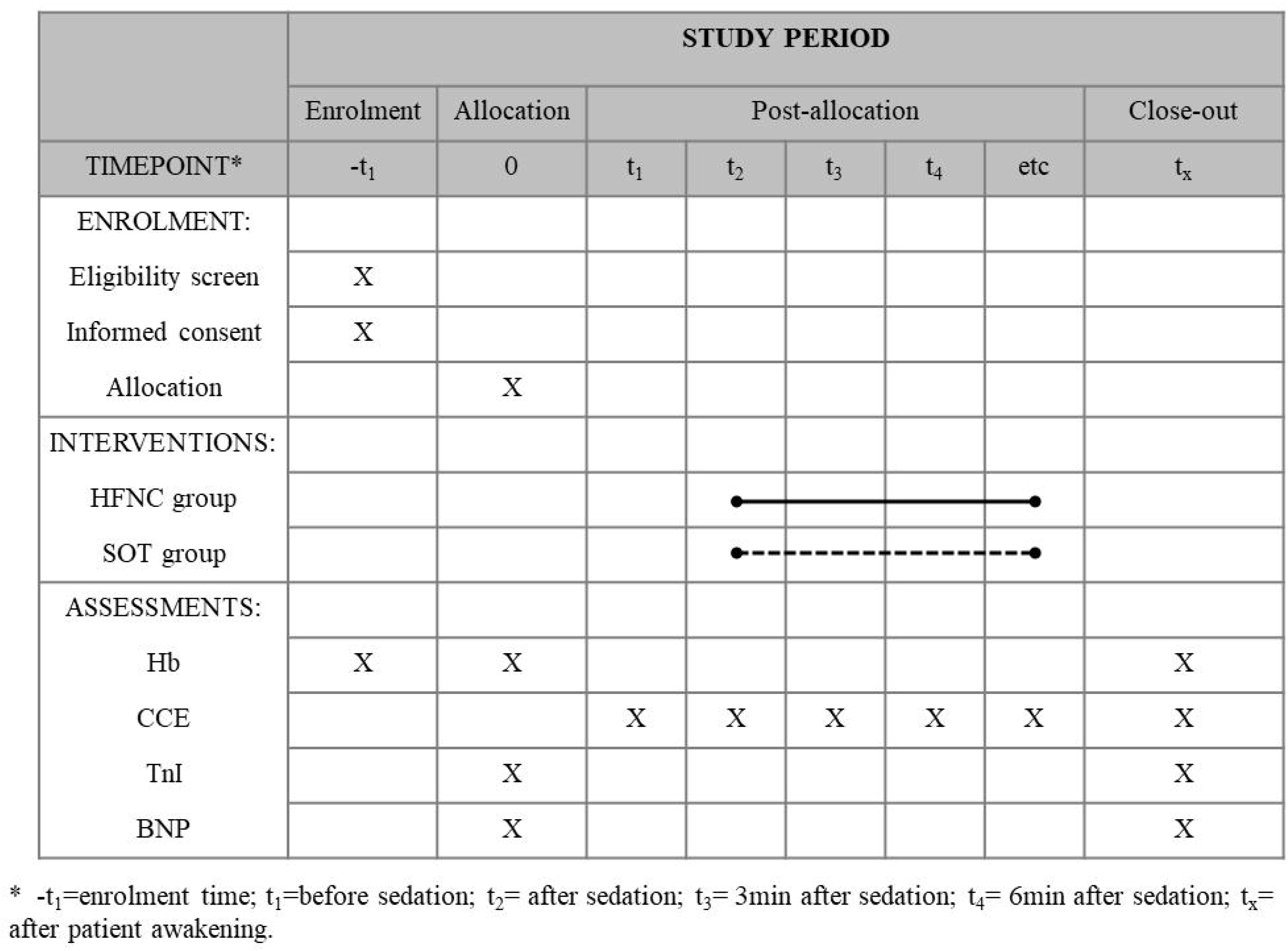
Example template for the schedule of enrolment, interventions, and assessments.

**Fig 2:**
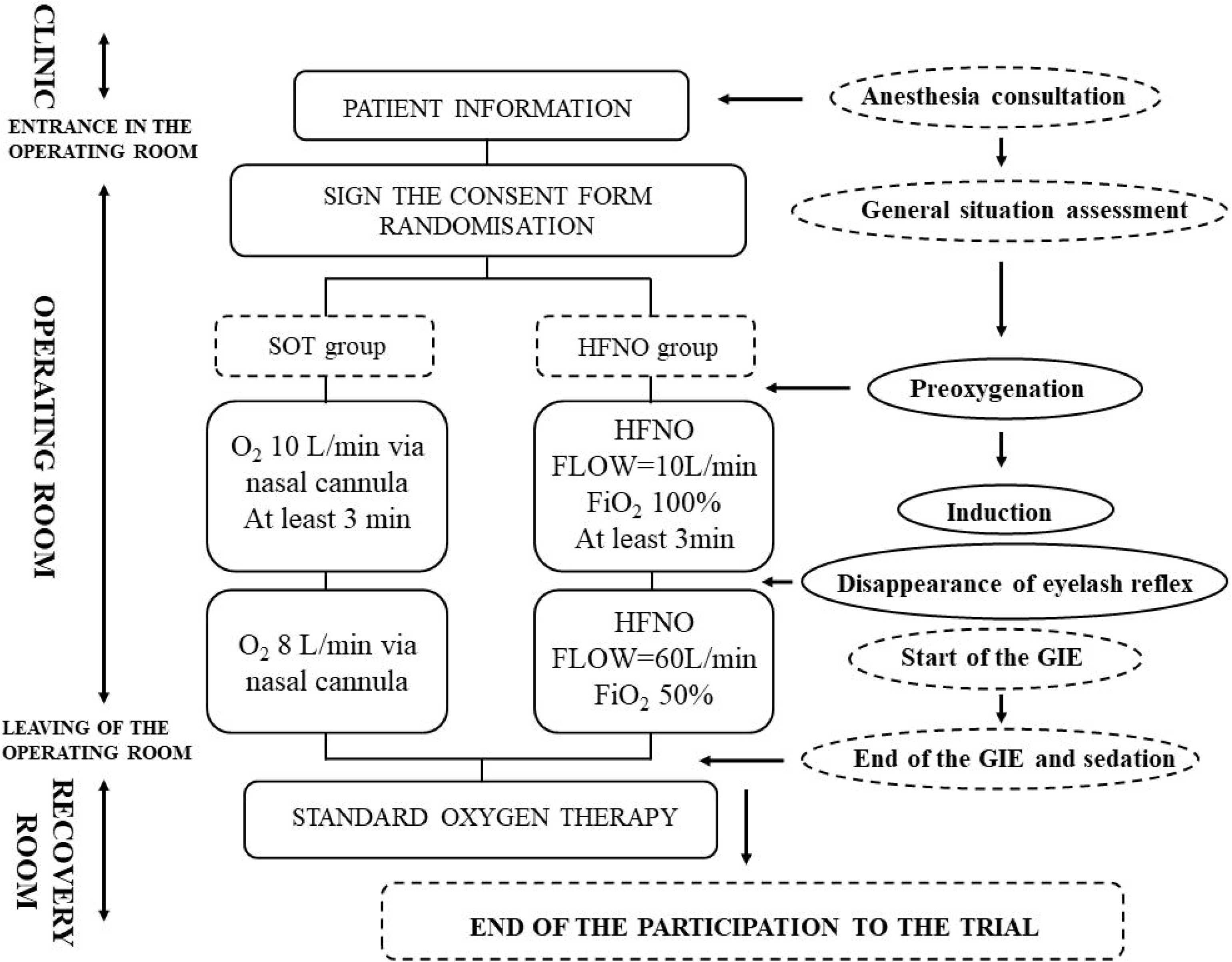
Flow of participants.

### Intervention

In the operating room, prior to commencing procedures, standard anesthesia monitoring equipment is used to monitor vital signs for each patient, including ECG, invasive blood pressure, and pulse oximetry. Additionally, hemodynamic data is monitored using the Mostcare monitor, and all data is recorded. Pulse oximetry is recorded in ambient air.

According to random allocation of interventions, preoxygenation is administered (low flow 10 L/min FiO2 100% for the HFNO group, 10 L/min for the SOT group) for at least 3 minutes prior to induction.

In this practical study, the investigators use sufentanil and propofol for anesthesia.

During anesthesia, the researchers administered oxygen therapy at 60 L/min in the HFNO group and at 8 L/min via nasal cannula in the SOT group. In cases of severe intolerance, HFNO or SOT may be discontinued and replaced by any other oxygen therapy technique (except for the SOT group). Tracheal intubation is permitted if necessary. In all instances, investigators must record all events in the case report form.

At the end of the GIE, patients are transferred to the recovery room, marking the end of the intervention period. In the recovery room, HFNO is not utilized. Instead, all patients are immediately administered SOT in the recovery room until it is deemed no longer necessary.

As in the study by Lin et al, hypercapnia is not monitored. To limit the risk of hypercapnia in our study, the FiO_2_ is set at 50% with a flow at 60□L/min.

### Intervention group

HFNO is administered using the HT-08 Optiflow Nasal High Flow device and dedicated anesthesia nasal cannula, equipped with filters (Aeonmed Medical System, China).

The preoxygenation settings entail a flow rate of 10 L/min on low-flow mode and 100% FiO2, sustained for at least 3 minutes. Induction follows after the preoxygenation period. Once the eyelash reflex disappears, switching to high-flow mode and the flow rate is increased to 60 L/min (to achieve a higher PEEP effect), and FiO2 is adjusted to 50%. The decision to set FiO2 at 50% is aimed at achieving similar FiO2 levels in both groups and reducing the risk of hyperoxia and hypercapnia.

### Control group

Oxygen therapy at a flow rate of 10 L/min via nasal cannula is administered for 3 minutes preoxygenation and 8 L/min throughout the entire anesthesia procedure.

### Similar FiO2

During the preoxygenation period, the FiO2 levels and flow rates are identical in both groups. In the HFNO group, FiO2 is set at 100% with a flow rate of 10 L/min in low-flow mode, while In the SOT group, the flow rate is set at 10 L/min with pure oxygen. This can provide equivalent and effective preoxygenation for both methods. For CAD patients under sedation with retained spontaneous breathing, pre-oxygenation can enhance tolerance during anesthesia.

During the procedure, we chose to set a similar initial FiO_2_ in both groups to prevent any disadvantage to the SOT group. Moreover, comparable FiO2 levels will aid in determining whether the positive end-expiratory pressure (PEEP) and dead space flushing effects induced by HFNO are beneficial. Similar FiO_2_ is obtained by using a conversion table. Therefore, after the standardized preoxygenation period, oxygen is administered to the SOT group via nasal cannula at a flow rate of 8 L/min to achieve a calculated FiO2 of 50%, while in the HFNO group, the oxygen flow will be set at 60 L/min with an FiO2 of 50%, ensuring equivalent FiO2 levels in both groups. Compared with an FiO2 of 100%, setting FiO2 at 50% in the HFNO group does not increase the incidence of hypoxemia.^36^

### Eligibility criteria

#### Inclusion criteria

- Patients older than 18 years old.
- Coronary heart disease patients scheduled for or requiring urgent GIE (upper and/or lower endoscopy), with plans for sedation aimed at maintaining spontaneous respiration (as determined during anesthesia consultation).
- Coronary heart disease patients are defined as those with single or multiple coronary artery stenosis >50% as determined by coronary angiography.

#### Exclusion criteria

- Necessity to intubate the patient for the procedure.
- Patient under oxygen therapy at home.
- Tracheostomised patient.
- High risk of aspiration, inability to fully clear upper respiratory secretions, thick or excessive airway secretions, and ineffective cough for sputum clearance.
- Hemodynamically unstable (sustained systolic arterial pressure <80 mmHg).
- Severe heart failure (ejection fraction <45%).
- Patients with severe cardiac arrhythmias (atrial fibrillation, frequent premature ventricular contractions, ventricular tachycardia, and second-degree or higher atrioventricular block).
- Unresolved tension pneumothorax or mediastinal emphysema.
- Combined with dysfunction of other organ systems (shock, gastrointestinal perforation/major bleeding, severe cerebral disorders, and severe hepatic or renal dysfunction).

### Outcome

The primary outcome is the difference in CEE levels between the two groups during the sedation. Secondary outcomes are the incidence of hypotension(hypotension is defined as a systolic blood pressure below 80 mmHg), values of BNP (brain natriuretic peptide), TnI (troponin I), and lactate levels at 6-12 hours post-operation,the occurrence of hypoxemia defined as SpO2 measurement equal to or below 92%, MostCare hemodynamic parameters excluding the primary outcome, interventions required to maintain upper airway patency, patient agitation episodes (assessed by touching the oxygen supply device), and presence of intraoperative adverse memories postoperatively.

### Statistical analysis

SPSS statistical software 23 (IBM-SPSS, Chicago, USA) will be used for data processing and analysis. The analysis report will adhere to the requirements of the Consolidated Standards of Reporting Trials statement.^37^ Raw data was tested for normality using the Kolmogorov-Smirnov and Bartlett tests.

The primary endpoint and the secondary hemodynamic parameters will be compared using distinct mixed linear models in which the patient will be considered as a random effect variable. The variable representing the randomized treatment group will be included as a fixed effect variable. The interaction between randomisation and stratification variables will be tested. The results will be presented in the form of estimated marginal means (and their 95% confidence intervals) for each randomized group. If the interaction is significant, results for each stratum group may also be presented. Secondary categorical endpoints will be compared between the two groups using either a χ2 test adjusted for stratification variables or student’s t-test.

The duration of sedation between the two groups will be compared using the Mann-Whitney U test, taking into account stratification. The interaction between the group of randomisation and the group of stratification will also be tested.

Subgroup analysis is planned: if there is a significant interaction between patient hemoglobin levels and randomized treatment groups, results will be presented stratified by hemoglobin levels. In case of missing data, multivariate imputation is planned. Missing values for the primary endpoint, CEE, will not be imputed or replaced.

### Ethics and dissemination

This study is a clinical research based on the premise of not compromising the rights and interests of the participants. The entire experiment strictly adheres to the current version of GCP, Chinese laws and regulations, and the management system of Anzhen Hospital for conducting clinical research. This ensures standardized procedures, scientifically reliable results, protection of participants’ rights, and assurance of their safety.

Paper medical record form was used as source document. The data will be entered into a secure spreadsheet application (Excel) by a member of the research team. The data will be handled in accordance with Chinese law. All original records will be archived at the test site for 30 years. Only four members of the MEHIS team (two first authors, the statistician, and the last author) will have access to the final experimental dataset.

In the event of an unexpected serious adverse event requiring review of the evolution of all enrolled patients, the Ethics Committee of Beijing Anzhen Hospital reserves the right to suspend or terminate the study at any time. Investigators may temporarily or permanently halt a patient’s participation for better patient service or in the event of a serious adverse reaction, and report the reasons.

The results will be submitted for publication in a peer-reviewed journal. The patient data from this clinical study will be treated as confidential and used only for the purposes of conducting this clinical research.

### Informed consent

Written consent (see consent form in online supplementary file 2) is obtained from all participants one day before performing the procedure.

After receiving appropriate disclosure of the potential risks and benefits of the study and having understood these explanations (see consent form in online supplementary file 1), participants were enrolled. Participant can contact the researchers at any time if they have any questions related to the study. By providing accurate contact information, they can contact the Ethics Committee of Anzhen Hospital at any time if they have any questions related to their own rights/interests. The participant is free to withdraw his/her consent to participate at any time.

## Discussion

The balance between myocardial oxygen supply and demand is a primary concern for anesthesiologists during invasive procedures (such as GIE performed under deep sedation) in spontaneous breathing patients with CAD. Assessment of CCE in patients with CAD based on MostCare data can effectively evaluate the oxygen supply and demand.

Due to the presence of multiple comorbidities in patients with CAD (such as anemia, hypertension and diabetes), achieving deep sedation while maintaining circulatory stability is crucial. While deep sedation in patients helps prevent stress reaction, maintaining the balance of myocardial oxygen supply and demand in sedated patients is crucial to ensure the safety of GIE procedures.

HFNO is widely used in sedation anesthesia for GIE. It is now gradually being extended to high-risk GIE patients to optimize oxygen delivery during anesthesia. To date, there is are few data regarding the use of HFNO during GIE in patients with CAD. Due to the low incidence of hypoxemia in the healthy population and the higher cost of HFNO compared to SOT, we chose to only include patients with CAD.

It is important that the flow rate and oxygen concentration during the pre-oxygenation phase were exactly the same for both groups. More importantly, the initial FiO2 plans are equivalent in both groups to avoid placing the control group at a disadvantage. Most studies compare HFNO with 100% FiO2 to standard oxygen at flow rates of 2-5 L/min, which roughly results in FiO2 between 28% and 45%.^38,39^ Contrarily, in the MEHIS trial, similar initial FiO2 in both groups will allow for the determination of whether the HFNO-induced PEEP and dead space washout effects are beneficial. Additionally, in the HFNO group, the gas flow is set to its higher value (60 L/min), contrary to previous studies, and all patients undergo preoxygenation before induction.

The study was launched on July 1, 2024. This rapid inclusion rate is expected to facilitate the acquisition of high-quality data by avoiding investigator and research team fatigue. So far, investigators have not reported any serious adverse events related to the study procedures. Dropouts for any reason are rare. All these factors inspire confidence in the timely completion of the study.

In summary, the MEHIS trial is a pragmatic randomized controlled trial initiated by investigators, aiming to test the hypothesis that HFNO, compared to SOT at similar FiO2 levels, can enhance CCE in patients with CAD undergoing GIE under deep sedation. The study presents several innovative aspects. Firstly, patients are at risk of myocardial oxygen imbalance. Secondly, the CCE of patients with CAD upper and/or lower GIE under deep sedation are investigated. Finally, the use of similar initial FiO2 levels aims to prevent the SOT group from being disadvantaged. If the results are positive, the use of HFNO could potentially become the standard approach for enhancing the safety of GIE performed under deep sedation in patients with CAD.

## Data Availability

All data produced in the present study are available upon reasonable request to the authors

## Data statement

Data set will be available on reasonable request to the corresponding author.

## Author statement

Sheng wang obtained funding. Fang Xie and Mu Jin designed the study. Tingting Ma and Xiaorui Zhou planned the statistical analysis. Sheng Wang and Fang Xie will have full access to the final trial data set. All authors participated in the writing the manuscript and approved the final version.

## Acknowledgments

The authors would like to thank the clinical staff of all trial sites (Beijing Anzhen Hospital).

## Ethics and dissemination

This study has been approved by the Ethics Committee of Beijing Anzhen Hospital, Capital Medical University (KS2024066), and patients are included after informed consent. The results will be submitted to a peer-reviewed journal for publication.

