## supplementary-material-1 for "MostCare-Based Assessment of Cardiac cycle efficiency in Coronary Artery Disease Patients: High-flow nasal cannula versus standard oxygenation for gastrointestinal endoscopy with sedation. The prospective single-center randomised controlled MEHIS study protocol"

**Keywords** high-flow nasal oxygen therapy (HFNO); cardiac cycle efficiency (CCE); gastrointestinal endoscopy (GIE); coronary artery disease（CAD）

**Trial registration number** ChiCTR2400086887

**Strengths and limitations of this study**

This is the first pragmatic randomized single-center study comparing HFNO to SOT for oxygenating patients with CAD undergoing GIE.

In contrast to comparing HFNO with high fraction of inspired oxygen (FiO_2_) typically used in SOT with low oxygen flow and consequently lower FiO2, this study adjusts gas flows to target roughly the same level of applied FiO2 in both groups. This approach aims to assess whether HFNO can improve CCE through positive end-expiratory pressure and/or dead space washout effects.

In SOT group, precise FiO2 cannot be guaranteed. That is why we utilized a pre-existing abacus to achieve the best equivalence.

Technically, blinding of practitioners and other nursing staff to the study groups is not feasible. However, the printout of recorded primary outcome measures is produced, allowing delayed reading by assessors blinded to treatment allocation. We employed a blind method in the study of patients.

In patients not undergoing endotracheal mechanical ventilation, MostCare data collection may exhibit slight bias due to respiratory influences. However, both groups in our study were conducted under sedation, and longitudinal comparisons were performed, thus minimizing the impact of such bias.

CAD patients with upper/lower gastrointestinal bleeding often suffer from anemia. We stratified CAD patients into anemic and non-anemic groups and compared the CCE under two oxygen delivery modes. This could provide a basis for further evaluating the oxygen therapy effectiveness of HFNO for anemic patients.

MostCare-Based Assessment of Cardiac cycle efficiency in Coronary Artery Disease Patients: High-flow nasal cannula versus standard oxygenation for gastrointestinal endoscopy with sedation. The prospective single-center randomised controlled MEHIS study protocol

Research Promoter: Beijing Anzhen Hospital, No. 2 Anzhen Road, chaoyang District, Beijing

Coordinating investigator: Dr Fang Xie, anesthesiology department, Beijing Anzhen Hospital, No. 2 Anzhen Road, chaoyang District, Beijing

Telephone number: +8601064456329

**Invitation**

You are invited by Doctor Fang Xie to participate in the clinical study titled " MostCare-Based Assessment of Cardiac cycle efficiency in Coronary Artery Disease Patients: High-flow nasal cannula versus standard oxygenation for gastrointestinal endoscopy with sedation. The prospective single-center randomised controlled MEHIS study protocol " at Anzhen Hospital in Beijing. Please carefully read this informed consent form and make a thoughtful decision on whether to participate in this study. You can ask your research doctor about anything related to this clinical research study that you do not understand. We encourage you to have a thorough discussion with your family and friends before making the decision to participate in this study. If you are participating in another study, please let your study doctor or research staff know. The purpose, background, research process and other important information of this study are given below.

**Background of this study**

High-flow nasal cannula oxygen therapy (HFNC) is a form of high-flow oxygen therapy in which patients are continuously provided with a constant concentration of inhaled oxygen and a constant temperature and humidity through a specialized, non-sealed nasal cannula. HFNO can provide higher oxygen flow rates and more precise fraction of inspired oxygen (FiO2), potentially offering better assessment of cardiac cycle efficiency (CEE) and myocardial protection. MostCare monitoring data of CCE is a comprehensive indicator reflecting the venous system, cardiopulmonary interaction, and coupling status between the ventricle and arterial system (V-A Coupling), which can effectively indicate whether the left ventricular function is optimized.

**What is the purpose of this study?**

The purpose is to investigate whether high-flow nasal oxygen therapy would decrease the incidence of myocardial ischemia by decreasing hypoxemia, circulatory fluctuations and improving CCE compared to standard oxygen therapy.

**How many people will participate in the study？**

Approximately 90 people (45 per group) will participate in this study. You will be placed in either SOT group (by nasal cannula with a flow at 8L/min) or in HFNC group（the high-flow nasal oxygen therapy with a flow at 60L/min）.

**What does this study involve？**

Coronary heart disease patients scheduled for or requiring urgent GIE (upper and/or lower endoscopy) with plans for sedation aimed at maintaining spontaneous respiration (as determined during anesthesia consultation) will be involved in. If you agree to participate in this study and sign the consent form, you will therefore be included in this study.

The main research objective is to compare the CCE. CCE is collected and generated by MostCare, recording values before and after sedation, every 3 minutes after induction, and after patient awakening. The secondary parameters are to compare between groups: the incidence of hypotension. Hypotension is defined as a systolic blood pressure below 80 mmHg; postoperative BNP, TnI, and lactate levels at 6-12 hours between the two groups; the incidence of hypoxemia (defined as SpO2 ≤92%); the frequency of patient agitation episodes (defined as touching the oxygen supply device); intraoperative adverse memory recall; the need for mask ventilation or any airway intervention; the duration of sedation (from the induction to the awakening of the patient). The secondary hemodynamic parameters are to compare between groups before and after sedation, every 3 minutes after induction, and after patient awakening: heart rate (HR); systolic blood pressure, diastolic blood pressure and mean arterial pressure, respectively (SBP, DBP and MAP); pdic-a; stroke volume index (SVI); systemic vascular resistance index (SVRI); cardiac index (CI); pulse pressure variation (PPV); the maximum rate of pressure increase in the left ventricle during systole (dP/dtmax).

**How long will this study continue?**

The study time includes the procedure of your painless gastroscopy and 12h after the procedure. You may opt out of the study at any time without penalty and without forfeiting any benefits you would otherwise receive. However, if along the way you decide to withdraw from this study, we encourage you to talk to your doctor first.

**What are the possible disadvantages?**

According to the literature, there are no risks associated with the use of high-flow nasal oxygen therapy. However, patients may experience discomfort due to the very high flow rate of oxygen. However, the treatment protocol calls for oxygen to be administered at a lower flow rate prior to sedation, and the flow rate increases once you are asleep. Therefore, you should not experience any discomfort.

**What are the benefits of participating in the study?**

If you agree to participate in this study, you will likely receive a direct medical benefit. High-flow nasal oxygen therapy may better maintain your intraoperative oxygenation and reduce the incidence of intraoperative hypoxia, while at the same time providing a greater supply of oxygen to your heart and protecting cardiac function. Mostcare machines are provided by the anesthesia department for free use by patients. We will use Mostcare to monitor your hemodynamic data in real time during surgery to provide you with safer anesthesia, and we hope that the information gained from your participation in this study will benefit patients with the same condition as you in the future.

**Will your information be kept confidential?**

With your understanding and assistance, the results of research conducted through this program may be published in medical journals. However, we will keep your research records confidential as required by law. Your personal information will be kept strictly confidential and will not be disclosed unless required by law. Government departments and hospital ethics committees and the relevant researchers may have access to your data as required.

**What are your legal rights?**

It is important that you understand the purpose of this study and the reasons why we are conducting it. If you decide not to take part in this study, it will not affect other treatments you are supposed to get. If you decide to participate, you will be asked to sign this written informed consent. You have the right to withdraw from the trial at any time during any stage of the trial without being discriminated against or treated unfairly, and your medical treatment and rights will not be affected.

If you have a serious adverse reaction, or if your study doctor feels that it is not in your best interest to continue participating in the study, he/she may decide to take you out of the study. The funder or regulatory agency may also terminate the study at any point during the study without your consent. We will also notify you in a timely manner if other conditions occur that affect your health, and your study doctor will discuss with you the other options you have.

**What are your responsibilities?**

You will be asked to provide truthful information about your own medical history and current physical condition and to tell the study doctor about any discomfort you have noticed during this study. If you have recently participated in other research studies or are currently participating in other research studies，you should also tell your study doctor.

**Who can you contact if you have questions or problems?**

If you have any questions related to this study, please contact Fang Xie at 18612510821 in the first instance. If you have any questions related to your rights/interests, or if you would like to reflect the difficulties, dissatisfaction and concerns encountered during your participation in this study, or if you would like to provide comments and suggestions related to this study, please contact the Ethics Committee of Beijing Anzhen Hospital Affiliated to Capital Medical University at 010-64456214.
