## supplementary-material-1 for "MostCare-Based Assessment of Cardiac cycle efficiency in Coronary Artery Disease Patients: High-flow nasal cannula versus standard oxygenation for gastrointestinal endoscopy with sedation. The prospective single-center randomised controlled MEHIS study protocol"

**Informational statement by the researcher**

I have informed the participant about the background, purpose, procedures, risks, and benefits of the study involving nasal high-flow oxygen therapy. I have given them enough time to read the informed consent form, discuss with others, and have answered their questions related to the research. I have informed the participant that they can contact Dr. Xie Fang at any time when they have questions related to the study. In case of issues related to their rights or interests, they can contact the Ethics Committee of Anzhen Hospital at any time, and I have provided accurate contact information. I have informed the participant that they have the option to withdraw from this study. I have also informed the participant that they will receive a copy of this informed consent form, which includes both my and their signature.

Signature of researcher	Date

**Subject Informed Consent Statement**

I have been informed about the background, purpose, procedures, risks, and benefits of the study on high-flow nasal oxygen therapy. I have enough time and opportunity to ask questions, and I am very satisfied with the answers provided. I have also been informed to contact whom when I have questions, want to report difficulties, concerns, suggestions for the study, or seek further information, or offer help for the research. I have read this informed consent form and agree to participate in this study. I am aware that I can withdraw from this study at any time during the research without providing any reason.

Signature of the subject	Date

Subject contact number

(When the participant's ability to give informed consent is lacking or insufficient, add or replace the following)

Signature of legal guardian	Date
